# The relative effectiveness of a high-dose quadrivalent influenza vaccine vs standard-dose quadrivalent influenza vaccines in older adults in France: a retrospective cohort study during the 2021-22 influenza season

**DOI:** 10.1101/2023.06.15.23291345

**Authors:** Bricout Hélène, Levant Marie-Cécile, Assi Nada, Crépey Pascal, Descamps Alexandre, Mari Karine, Gaillat Jacques, Gavazzi Gaétan, Grenier Benjamin, Launay Odile, Mosnier Anne, Raguideau Fanny, Watier Laurence, Rebecca C Harris, Chit Ayman

## Abstract

**Background:** High-dose quadrivalent influenza vaccine (HD-QIV) was introduced during the 2021/22 influenza season in France for adults aged ≥65 years as an alternative to standard-dose quadrivalent influenza vaccines (SD-QIV). This is the first study to estimate the relative vaccine effectiveness (rVE) of HD-QIV versus SD-QIV against influenza-related hospitalizations in France.

**Methods:** Community-dwelling individuals aged ≥65 years with reimbursed influenza vaccine claims during the 2021/22 influenza season were included from the French national health insurance database. Individuals were followed up from vaccination day to 30 June 2022, nursing home admission or death date. Baseline socio-demographic and health characteristics were identified from medical records over the 5 previous years. Hospitalizations due to influenza and other causes were recorded from 14 days after vaccination to end of follow-up. HD-QIV and SD-QIV vaccinees were matched using 1:4 propensity score matching with an exact constraint on age group, sex, week of vaccination and region. Incidence rate ratios (IRR) were estimated using zero-inflated Poisson or zero-inflated negative binomial regression models.

**Results:** We matched 405,385 (99.9%) HD-QIV to 1,621,540 SD-QIV vaccinees. HD-QIV was associated with a 23.3% (95%CI: 8.4–35.8) lower rate of influenza hospitalizations compared to SD-QIV. Post-matching, we observed higher rates in the HD-QIV group for hospitalizations non-specific to influenza and for negative control outcomes, suggesting residual confounding by indication.

**Conclusions:** HD-QIV was associated with lower influenza-related hospitalization rates versus SD-QIV, consistent with existing evidence, in the context of high SARS-CoV-2 circulation in France and likely prioritization of HD-QIV for older/more comorbid individuals.

## Introduction

Influenza has a significant public health impact. In France, over the past 10 seasons (excluding 2020-21), influenza infection was responsible for more than 1 million general practitioners (GP) visits for Influenza-Like Illness (ILI), along with more than 20,000 influenza hospitalizations and around 9,000 deaths each season (1). These surveillance figures likely underestimate the true burden of seasonal influenza. The French Agency of Public Health, Santé Publique France, reported that 90% of influenza-related deaths occur in patients aged 65 years and over(2). Complications leading to hospitalizations and deaths are greatest among persons aged 65 years and older since they are often more vulnerable due to chronic disease and weakened immune systems. Thus, with the overall aim of reducing the public health burden of influenza, in particular the associated hospitalizations, annual influenza vaccination is recommended and free of charge in France for this age group.

Standard-dose (SD) influenza vaccines are considered to provide suboptimal protection for adults over the age of 65 years (3), due to an age-related decline in immune system function (immunosenescence) associated with a decrease in the response to vaccination (4). A high-dose (HD) influenza vaccine, with higher antigen content than the SD vaccine, has been developed to elicit an improved immune response and improved protection against severe influenza illness, influenza-related hospitalizations, and mortality in this vulnerable population. In a pivotal randomized controlled trial, HD influenza vaccine demonstrated a superior relative vaccine efficacy of 24.2% (95%CI: 9.7;36.5%) vs SD influenza vaccine in preventing laboratory-confirmed influenza (5). Furthermore, a recent meta-analysis of published evidence of HD vs SD vaccine efficacy/effectiveness studies showed that HD was associated with 11.7% (95% CI: 7.0-16.1%) reduction in influenza related hospitalizations, 27.3% (95% CI: 15.3-37.6%) reduction in pneumonia hospitalizations, 13.4% (95% CI: 7.3-19.2%) reduction in combined pneumonia/influenza hospitalizations and 17.9% (95% CI: 15.0-20.8%) reduction in cardiorespiratory hospitalizations (6).

Following a decade of use of HD trivalent influenza vaccine (HD-TIV), mainly in the US, HD quadrivalent influenza vaccine (QIV), containing antigens from two influenza A strains and both B lineage strains, was introduced from season 2020/21 in the US and from season 2021/22 in Europe. This study estimated the relative vaccine effectiveness (rVE) of HD-QIV vs SD-QIV against hospitalization outcomes in community-dwelling adults 65 years of age and older during the 2021/22 influenza season in a real-world setting in France.

## Methods

This was an observational retrospective cohort study, based on national administrative healthcare data from France, designed to describe the characteristics of individuals who received a seasonal influenza vaccine in the community between September 1st, 2021, and February 28, 2022, and to assess the rVE of HD-QIV compared with SD-QIV for hospitalization outcomes. The study used data from the National Health Data System (Système National Des Données de Santé; SNDS), part of the National Health Insurance (NHI) system. The SNDS collates health data collected by NHI agencies. We used data from two pre-existing databases: the NHI Système National d’Information Inter-Régimes de l’Assurance Maladie (SNIIRAM) and the hospital discharge database (Programme de Médicalisation des Systèmes d’Information; PMSI). The SNDS contains individual-level data for outpatient and private healthcare facilities health expenditure billing for reimbursement purposes from the SNIIRAM that are linked to the hospitalization database PMSI using a unique, anonymous identifier, the social security number (Numéro d’inscription au répertoire de l’INSEE; NIR). As such, it encompasses anonymous, individual-level data for all healthcare claims for more than 99% of the population residing in France, regardless of the insurance scheme, i.e., close to 65 million people (7–10).

Individuals included in the study were 65 years of age or older on the day of vaccination with a unique social security number, together with a record of influenza vaccine dispensed at a pharmacy between September 1^st^, 2021, and February 28, 2022 (official end of the French influenza vaccination campaign). We used pharmacy dispense records as a proxy for influenza vaccination. In the French health care system, individuals who are eligible for influenza vaccination receive a voucher for the vaccine to be dispensed at a pharmacy and then administered by a pharmacist, general practitioner (GP) or nurse (as GP surgeries in France do not store vaccines themselves). We assumed the date of vaccination (“index date”) was approximated by the date of influenza vaccine pharmacy dispensation. Vaccine type was classified by medication codes, as listed in Supplementary Table S1. Individuals were excluded if they experienced an outcome related to hospitalization from the start of the influenza season up to 14 days after the index date. Individuals receiving multiple influenza vaccines in the same season, those who were vaccinated with a Northern Hemisphere (NH) 20/21 formulation and individuals for whom information on region and/or deprivation index were not available were also excluded.

A fixed 5-year pre-index date period (September 1^st^, 2016, to June 30^th^, 2022) was used to capture baseline demographics, comorbidities/medical history, and previous treatments and vaccinations. The same period is usually used by the NHI to identify a specified set of pathologies (11) . Individuals were in the cohort from the date of vaccination up to 30 June 2022 or up to admission to medico-social housing or a nursing home, or death. Hospitalizations due to influenza and other causes were collected from 14 days after the index date (start of vaccine protection) to end of follow-up. Primary outcomes were hospitalizations for influenza, pneumonia, respiratory disease, cardiovascular disease, and cardiorespiratory (either cardiovascular or respiratory) disease. Hospitalizations were ascertained by their ICD-10 discharge diagnosis code (Supplementary Table S2). Hospitalizations with a COVID-19-associated discharge diagnosis code were excluded from this primary analysis. Given that the PMSI hospital administrative database is maintained for reimbursement purposes, we used both primary ICD-10 discharge diagnosis codes and non-primary diagnosis codes to identify the study outcomes for the analysis. Given the primary diagnosis code should be the main determinant for hospitalization (12), we reported primary analyses based on the primary ICD-10 discharge code. However, as the choice of coding in PMSI could potentially be impacted by the level of severity of the outcome and linked to that, by the level of reimbursement that can be claimed by the hospital, we also conducted an analysis with the outcomes of interest coded with both primary or non-primary discharge codes.

Since the study relied on retrospective database analysis with routine treatment allocation (no randomization), a treatment selection or indication bias could exist. To adjust for potential confounding, every HD-QIV vaccinee was matched to four SD-QIV vaccinees. Propensity score matching was performed using a greedy matching algorithm without replacement with a caliper value equal to 20% of the variability (standard deviation) of the logarithm of the propensity score values (0.2*STD(log(PS)), where PS=propensity score, with an exact matching constraint on sex, age group (65 to 75 years of age, 75 to 85 years of age, and over 85 years of age), geographical region and week of dispensing of the vaccine (± 1 week). Geographical region and week of dispensing were included to address temporal and geographical factors likely associated with access to HD-QIV (e.g., supply) and influenza virus exposure (i.e., viral activity and circulation). The propensity score was computed using a logistic regression model with vaccine group as the dependent variable and all relevant covariates as independent variables. These independent variables included: number of GP consultations in the past 12 months, number of hospitalizations in the past 12 months, number of influenza vaccinations (in the 5 previous seasons), age (continuous, categories: below 75 years, 75-85 years, above 85 years), sex, vaccine administration setting (either at pharmacy or not), quintiles of the French social deprivation index (FDep), vaccination status for influenza in the preceding season (HD-QIV, SD-QIV, not vaccinated), COVID-19 vaccination status (completed, on-going [i.e. partial], unvaccinated), pneumonia vaccination status in the past 5 years, geographical region, and a diagnosis in the previous 5 years for diabetes, obesity and/or history of obesity surgery (referred to hereon as “obesity”), undernourishment or history of undernourishment (referred to hereon as “malnutrition”), chronic obstructive pulmonary disease (COPD)/asthma, dementia, myocardial infarction, chronic coronary disease, chronic heart failure, peripheral artery disease (PAD), hematological tumors, solid tumors, solid organ transplant, stem cells transplant, HIV (human immunodeficiency virus), drug-induced immunosuppression, chronic liver diseases and severe renal disease, and number of comorbidities of interest. The ICD-10 codes used to identify each of the comorbidities listed are described in Supplementary Table S3.

To estimate the association between vaccination with HD-QIV or SD-QIV and study hospitalization outcomes, a zero-inflated Poisson or zero-inflated Negative binomial regression model(13) was used to estimate incidence rate ratios (IRR) along with corresponding 95% confidence intervals (CIs). The choice of the regression model method was based upon the Akaike information criterion A (14). The models included an offset for the log of the follow-up time which allows a rate model to be computed (equivalent to specifying the outcome as the number of hospitalizations divided by the length of the follow-up period). The rVE was computed as ([1-RR] * 100%), along with their corresponding 95% CIs by Taylor series variance approximation (15, 16).

We performed a number of sensitivity analyses. For the first of these, we restricted outcomes to those occurring within a 9-week interval encompassing the 4 weeks before and the 4 weeks after the week of peak incidence (as defined by Santé Publique France). This interval coincided with weeks 9 to week 17 of the study, inclusive (28 February to 1 May 2022) (17, 18). This analysis was designed to increase the specificity of the hospitalization outcomes of interest, as the probability of miscoding influenza hospitalizations should be lower during peak influenza activity; and the probability of being hospitalized for respiratory/cardiovascular events due to influenza should be higher. We also explored the impact of using different outcome definitions (primary discharge code vs. any primary or non-primary discharge diagnosis code), and individuals with outcomes of interest that also included a COVID-19 discharge ICD-10 code. Indeed, SARS-CoV2 was circulating and was under a high level of surveillance, thus COVID-19 infection may have been frequently coded and may have played a role in some of the hospitalizations, especially in hospitalizations that were non-specific to influenza. Finally, we conducted negative control outcomes (falsification) analysis to check for unmeasured confounding by examining the effect of HD-QIV versus SD-QIV on hospitalizations that were unrelated to influenza, to influenza complications or to influenza vaccination (i.e., urinary tract infection [UTI], cataract surgery, or erysipelas).

## Results

We identified 8,136,621 individuals aged 65 years or older who received an influenza vaccine in France during the 2021/22 influenza season (Figure 1), of which 303,368 were excluded as they received vaccination in a nursing home setting. We excluded a further 25,908 and 409,997 HD-QIV and SD-QIV vaccinees, respectively, due to receipt of more than one influenza vaccine, missing data on region or deprivation index, or experiencing outcomes of interest during the period between the beginning of the influenza season and up to two weeks after the date of vaccination. We were then able to match 405,385 HD-QIV vaccinees to 1,621,540 SD-QIV vaccinees, achieving a 1:4 match ratio for these 2,026,925 included individuals. The 350 HD-QIV vaccinees that did not achieve a 1:4 matching ratio (0.08%) were excluded from the analysis and their characteristics are summarized in the supplemental materials (Supplementary Table S4).

**Figure 1:**
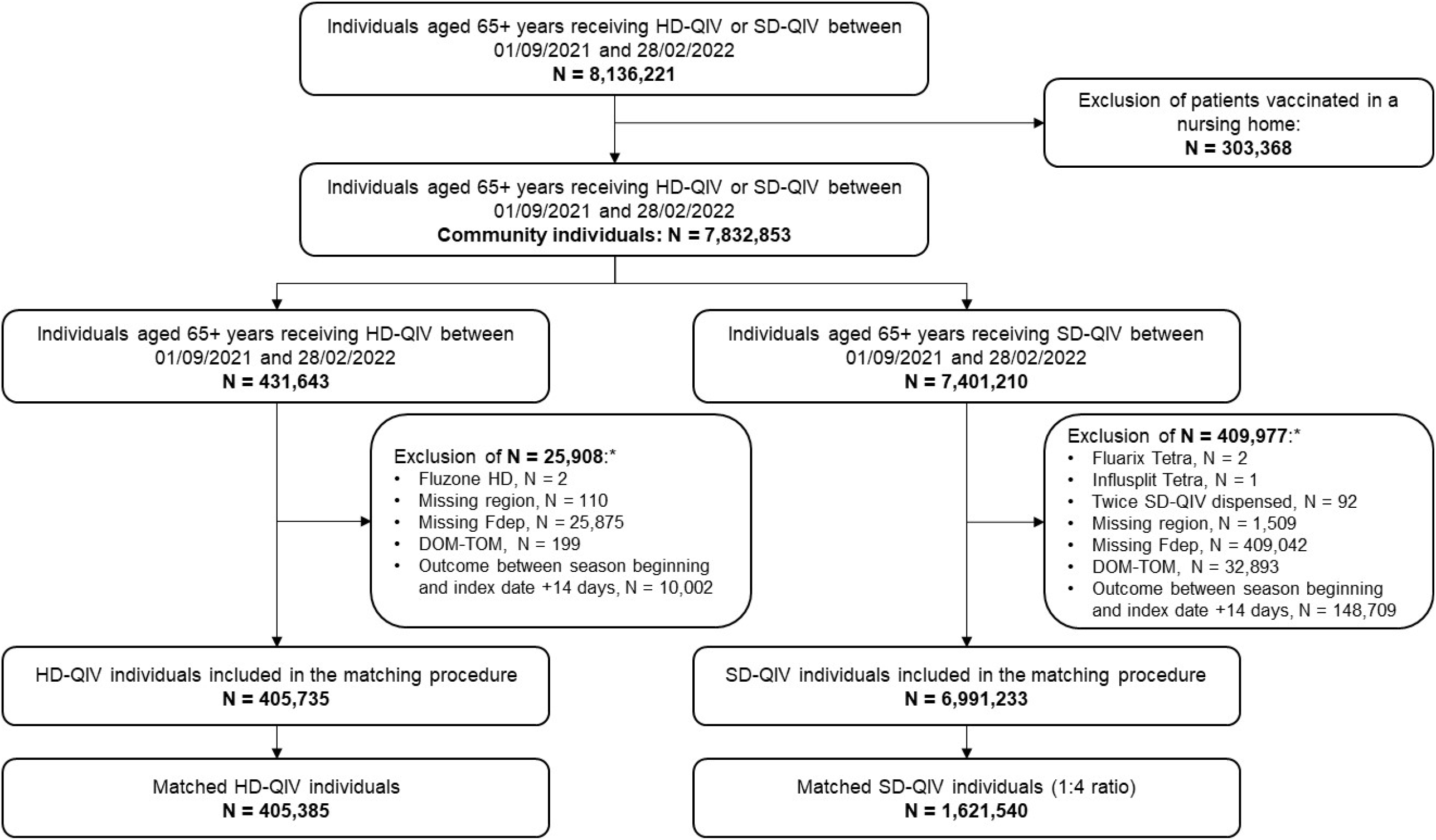
Flowchart for community SD-QIV and HD-QIV recipients aged 65 years and older during the 2021/2022 season in France. * More than one exclusion criterion may have been applied to a single patient, e.g. an excluded patient may both have a missing Fdep and be residing in the DOM-TOM (French overseas departments and territories). DOM-TOM, French overseas departments and territories; Fdep, French social deprivation index; HD, high-dose; QIV, quadrivalent influenza vaccine; SD, standard dose;

Baseline characteristics of the HD-QIV and SD-QIV cohorts before matching and after matching are described in Supplementary Table S5 and Table 1. At baseline, HD-QIV recipients were older than SD-QIV recipients (mean age, 77.4 vs 75.9 years; p<.0001), and had a significantly higher prevalence of most comorbidities of interest and of multiple comorbidities (54.8% of HD-QIV vaccinees had at least 1 comorbidity versus 52.4% for SD-QIV; p<.0001). During the follow-up period, HD-QIV recipients also experienced more severe outcomes post-hospitalization (death rates of 2.0% vs 1.5%; p<.0001). After matching, characteristics were similar between the cohorts. However, there was a slight trend towards a marginally higher prevalence of any chronic diseases (for example, 27.9% HD-QIV individuals with history of cardiovascular diseases vs 26.7% for SD-QIV), prevalence of multiple chronic diseases (55.0% with at least 1 comorbidity vs 51.8%) and death rates in the HD-QIV group (1.9% vs 1.6%).

**Table 1:** Baseline cohort characteristics – after matching

| Characteristics | HD-QIV | SD-QIV |
| --- | --- | --- |
| Number of individuals | 405,385 | 1,621,540 |
| Age, mean (± STD) | 77.4 (7.4) | 77.3 (7.9) |
| • 65-75 years, n (%) | 172,071 (42.4) | 688,284 (42.4) |
| • 75-85 years, n (%) | 161,258 (39.8) | 645,032 (39.8) |
| • Over 85 years of age, n (%) | 72,056 (17.8) | 288,224 (17.8) |
| Women, n (%) | 227,755 (56.2) | 911,020 (56.2) |
| *Reasons for end of follow up, n (%)* |  |  |
| Admission into a medico-social housing (other than NH) | 35 (0.0) | 121 (0.0) |
| Admission into NH | 1,416 (0.3) | 5,178 (0.3) |
| Death | 7,605 (1.9) | 26,608 (1.6) |
| End of follow-up | 396,329 (97.8) | 1,589,633 (98.0) |
| *Health care seeking behaviors proxy* |  |  |
| All-cause hospitalization in the past 12 months, mean (STD) | 0.1 (0.9) | 0.1 (0.8) |
| GP visits in the past 12 months, mean (STD) | 6.2 (4.8) | 6.1 (4.6) |
| Influenza vaccination at pharmacy, n (%) | 205,005 (50.6) | 834,196 (51.4) |
| Influenza vaccination during the previous season, n (%) | 369,734 (91.2) | 1,490,257 (91.9) |
| COVID-19 vaccinated*, n (%) | 391,967 (96.7) | 1,571,018 (96.9) |
| Pneumococcal vaccination in the previous 5 years, n (%) | 47,432 (11.7) | 179,405 (11.1) |
| *Medical conditions during the 5 years prior index date, n (%)* |  |  |
| Diabetes | 80,454 (19.9) | 309,552 (19.1) |
| Obesity and/or history of obesity surgery | 34,503 (8.5) | 129,287 (8.0) |
| Undernourishment/or history of undernourishment | 27,848 (6.9) | 100,485 (6.2) |
| COPD/Asthma | 48,102 (11.9) | 177,592 (11.0) |
| Dementia | 13,607 (3.4) | 46,906 (2.9) |
| Cardiovascular diseases | 113,066 (27.9) | 432,428 (26.7) |
| Immunocompromised individuals | 75,142 (18.5) | 285,269 (17.6) |
| Chronic liver disease | 6,525 (1.6) | 24,125 (1.5) |
| Terminal chronic kidney failure | 1,692 (0.4) | 6,303 (0.4) |
| *Number of chronic diseases, n (%)* |  |  |
| None | 182,213 (45.0) | 781,010 (48.2) |
| 1 | 130,086 (32.1) | 494,610 (30.5) |
| 2 | 59,146 (14.6) | 218,801 (13.5) |
| 3 | 22,584 (5.6) | 84,959 (5.2) |
| 4 | 7,981 (2.0) | 29,673 (1.8) |
| 5 | 2,536 (0.6) | 9,206 (0.6) |
| 6 | 839 (0.2) | 3,281 (0.2) |
| *Precariousness index (FDep99), n (%)* |  |  |
| Q1, Individuals living in a low-poverty municipality | 77,989 (19.2) | 313,265 (19.3) |
| Q2 | 79,443 (19.6) | 318,652 (19.7) |
| Q3 | 85,160 (21.0) | 340,820 (21.0) |
| Q4 | 79,318 (19.6) | 315,693 (19.5) |
| Q5, Individuals living in a highly disadvantaged municipality | 83,475 (20.6) | 333,110 (20.5) |

\*COVID-19 vaccinated is a variable identified as such within the database. It reflects the COVID-19 vaccination status of each patient at index date following current guidelines (it can refer to a single dose, two, or three, depending on the individual's eligibility)
Standard differences showed good balance for all variables included in the matching procedure (i.e. Absolute value of std. diff < 0.1)

During the study period, the hospitalization rates with influenza as the primary ICD-10 discharge code were 69.5 per 100,000 person years and 90.5 per 100,000 person years for HD-QIV and SD-QIV recipients, respectively, translating to an rVE of 23.3% (95% CI: 8.4–35.8) (Table 2). In the sensitivity analysis restricting outcomes to the peak influenza period, influenza hospitalization rates were 52.6 per 100,000 person years and 72.4 per 100,000 person years for HD-QIV and SD-QIV, respectively, yielding a point estimate rVE of 27.4% (95% CI: 11.1–40.7). In the sensitivity analysis including influenza hospitalizations with a COVID-19 code, influenza hospitalization rates for HD-QIV and SD-QIV were 70.3 per 100,000 person years and 92.0 per 100,000 person years, respectively, yielding an rVE of 23.6% (95% CI: 8.9–36.0), similar to the main analysis.

**Table 2:** Relative vaccine effectiveness and sensitivity analysis against influenza hospitalizations

| **Vaccine group** | **Hospitalization rate**<br>**per 100,000 person years (95% CI)** | **rVE (95% CI)** | **P-value** |
| --- | --- | --- | --- |
| HD QIV | 69.47 (59.64-80.92) | 23.29 [8.38;35.77] | 0.003 |
| SD QIV | 90.53 (84.68-96.78) |  |  |
| *Sensitivity analysis including influenza hospitalizations with a COVID-19 code* |  |  |  |
| HD QIV | 70.31 (60.42-81.83) | 23.61 [8.88;35.96] | 0.003 |
| SD QIV | 92.00 (86.10-98.29) |  |  |
| *Sensitivity analysis during the peak of the season* |  |  |  |
| HD QIV | 52.63 (44.17-62.71) | 27.38 [11.05;40.70] | 0.002 |
| SD QIV | 72.36 (67.15-77.97) |  |  |

For the hospitalization outcomes that were non-specific to influenza, we observed marginally higher hospitalization rates in HD-QIV than SD-QIV recipients (Table 3). In the sensitivity analysis of these non-influenza-specific hospitalization outcomes, restricted to the period of peak influenza activity, the rates for pneumonia hospitalization, pneumonia and/or influenza hospitalization, respiratory hospitalization and cardiorespiratory hospitalization tended to be slightly lower for HD-QIV than for SD-QIV (IRR ≤1) (Table 4). When alternative outcome definitions were used, using primary or non-primary diagnosis, similar results were observed, with an IRR of 0.79 (95% CI: 0.68–0.91) for influenza hospitalizations, and IRRs slightly superior to 1 for the non-influenza-specific hospitalization outcomes (Supplementary Table S6). The sensitivity analysis of non-influenza-specific hospitalizations including outcomes with a COVID-19 code showed IRR slightly above 1 (Supplementary Table S7).

**Table 3:** Non-influenza specific post vaccination outcome rates identified in primary diagnosis, excluding COVID-19 codes

| **Hospitalization**<br>**outcomes** | **HD-QIV event rate Per**<br>**100,000 person-years** | **SD-QIV event rate Per**<br>**100,000 person-years** | **IRR HD-QIV vs SD-**<br>**QIV (95% CI)** |
| --- | --- | --- | --- |
| Pneumonia | 701.02 (668.15-735.52) | 681.99 (665.61-698.78) | 1.03 (0.97;1.09) |
| Pneumonia and<br>influenza | 770.49 (735.99-806.61) | 772.52 (755.07-790.38) | 1.00 (0.94;1.06) |
| Respiratory | 942.69 (904.45-982.56) | 922.91 (903.81-942.41) | 1.02 (0.97;1.08) |
| Cardiovascular | 4,078.98 (3,998.56-4,161.01) | 3,966.88 (3,927.08-4,007.09) | 1.03 (1.00;1.06) |
| Cardiorespiratory | 4,858.74 (4,770.90-4,948.20) | 4,750.02 (4,706.45-4,793.99) | 1.02 (1.00;1.05) |

**Table 4:** Sensitivity analysis for non-influenza-specific hospitalization outcomes during the peak influenza season

| **Hospitalization outcomes** | **HD-QIV event rate Per**<br>**100,000 person-years** | **SD-QIV event rate Per**<br>**100,000 person-years** | **IRR HD-QIV vs SD-QIV**<br>**(95% CI)** |
| --- | --- | --- | --- |
| Pneumonia | 190.31 | 191.34 | 1.00 (0.89; 1.11) |
| Pneumonia and influenza | 242.94 | 263.70 | 0.92 (0.84; 1.02) |
| Respiratory | 294.72 | 311.91 | 0.95 (0.87; 1.03) |
| Cardiovascular | 1,191.53 | 1,185.04 | 1.01 (0.96; 1.05) |
| Cardiorespiratory | 1,437.41 | 1,451.57 | 0.99 (0.95; 1.03) |

Negative control outcomes analysis showed IRRs of 1.03 (95% CI: 0.98–1.07) against UTI hospitalizations, 1.00 (95% CI: 0.98–1.02) against cataract hospitalizations and 1.05 (95% CI: 0.96–1.16) against erysipelas hospitalizations. Although none were statistically significant, there was a slight trend towards higher rates of hospitalizations in HD-QIV vs SD-QIV recipients for the three different falsification outcomes considered (Table 5), which may suggest residual unmeasured confounders.

**Table 5:**
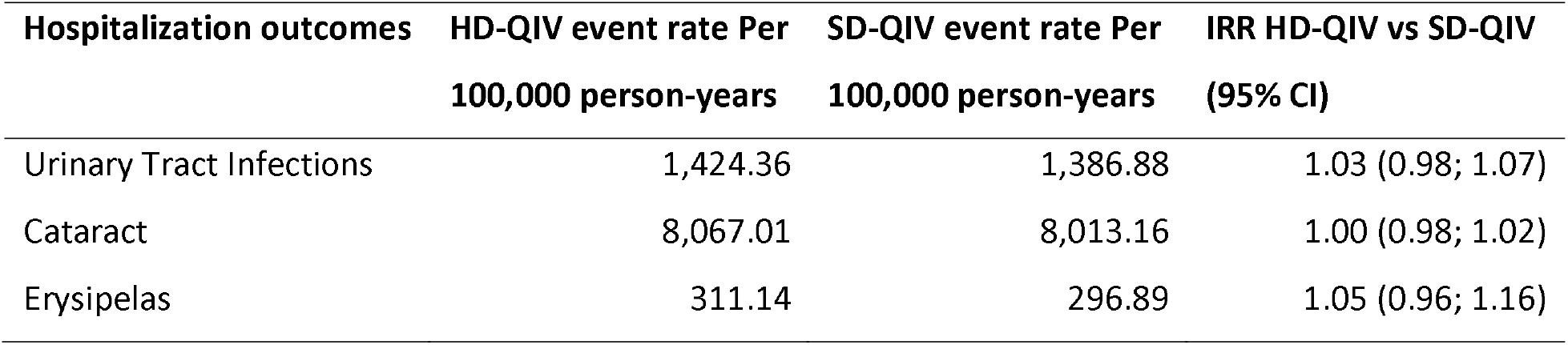
Falsification analysis results

| **Hospitalization outcomes** | **HD-QIV event rate Per**<br>**100,000 person-years** | **SD-QIV event rate Per**<br>**100,000 person-years** | **IRR HD-QIV vs SD-QIV**<br>**(95% CI)** |
| --- | --- | --- | --- |
| Urinary Tract Infections | 1,424.36 | 1,386.88 | 1.03 (0.98; 1.07) |
| Cataract | 8,067.01 | 8,013.16 | 1.00 (0.98; 1.02) |
| Erysipelas | 311.14 | 296.89 | 1.05 (0.96; 1.16) |

## Discussion

In this study, we assessed vaccine effectiveness of HD-QIV vs SD-QIV against influenza hospitalisations for the first time in France, in 405,385 HD-QIV recipients aged 65 years and older matched to 1,621,540 SD-QIV recipients. HD-QIV was associated with 23.3% (95% CI: 8.4–35.8) fewer hospital admissions due to influenza compared to SD-QIV, a finding that was consistent across sensitivity analyses. This finding is aligned with the literature, with a rVE against influenza hospitalization of HD vs SD influenza vaccines estimated at 11.7% (95% CI: 7.0–16.1) in a recently updated meta-analysis,(6) and a large Danish pragmatic RCT in 2021/22 with an observed rVE of 64.4% (95% CI: 24.4-84.6) for hospitalizations for pneumonia and/or influenza in HD-QIV recipients vs SD-QIV recipients (19). No differences or non-significant trends in favor of HD-QIV vs SD-QIV were observed for hospitalization outcomes that were non-specific to influenza (defined with primary ICD-10 discharge codes). We also observed a non-significant trend towards higher rates of negative control outcomes in HD-QIV recipients, suggesting that, despite matching on several measurable baseline characteristics, residual confounding by indication (i.e. in the case of HD-QIV being preferentially administered to more vulnerable individuals and individuals with higher levels of comorbidity) and residual unmeasured confounding may have remained. Indeed, while the two cohorts were matched based on a number of baseline characteristics, we still observed a slight trend towards marginally higher prevalence of chronic diseases. Despite this potential residual confounding biased against the HD-QIV vaccine, rVE against influenza hospitalization was still observed, and was robust to the sensitivity analyses run. Furthermore, our findings support previously published data showing improved protection provided by HD-QIV compared with SD-QIV in elderly individuals in a randomized setting (5).

Limitations of our study included potential bias in the distribution of the HD-QIV vaccine during the first year of its release in France. Indeed, the French Geriatric Scientific Society (Société Française de Gériatrie et Gérontologie, SFGG) recommends the preferential use of HD-QIV to all adults aged 65 years of age and over;(20) however, to support supply management during this first year following its release, the SFGG recommended prioritization of HD-QIV for at-risk seniors, i.e. older and/or with multiple comorbidities for the first year of use (1). The slightly higher hospitalization rates for negative control outcomes and for some non-influenza-specific outcomes (based on primary and non-primary discharge codes) in the HD-QIV recipients in our study supported this 2021/22 recommendation being implemented in practice, as did baseline characteristics before matching (HD-QIV vaccinees were older and had more co-morbidities than SD-QIV vaccinees). The limitations linked to the use of administrative databases for research purposes also applied to our study. Indeed, we may not have captured all relevant health characteristics of the study individuals. This limits the ability of our matching techniques to adjust for confounding, as without robust estimators of frailty or level of severity for the comorbidity, it is difficult to adjust for these confounding factors, which may lead to an underestimation of vaccine effectiveness. Other unmeasured confounders missing from administrative databases could have impacted our findings if they were unbalanced between HD-QIV and SD-QIV recipients, including for example frequency of contact with children and the proportion living alone (social isolation). In addition, while we used the pharmacy dispensing date as a proxy for vaccination date, around 50% of those in the study were not vaccinated at the pharmacy (no pharmacy vaccination date recorded). However, sensitivity analyses using first medical encounter within 2 weeks of pharmacy collection instead of pharmacy dispensing data did not alter the study findings (data not shown). Finally, atypical viral epidemiology (late influenza peak in April 2022, moderate intensity influenza epidemic and SARS-CoV-2 co-circulation) (21) in the 2021/22 season may have also contributed to the low to no effect observed for non influenza-specific hospitalization outcomes. In this context of a moderate influenza season, influenza contributes less to these broader outcomes (e.g., pneumonia, respiratory or cardiovascular hospitalizations), therefore diluting the effect of influenza vaccines in preventing those potentially influenza-associated complications, leading to potential underestimation of rVE.

The strengths of our study included the large sample size and our ability to capture all HD-QIV vaccines provided free of charge at pharmacies in the community setting in France, giving a comprehensive overview of vaccine effectiveness. We set the level of statistical significance for our analysis at 5%, as commonly used in the scientific literature, but considering the very large sample size, we should interpret with caution any small effect sizes found to be significant (22, 23). Additionally, increased use of PCR testing for influenza and other respiratory viruses during the COVID-19 pandemic (24, 25) was likely to have improved the specificity of influenza coding at hospital discharge, allowing for more accurate identification of influenza cases for the influenza hospitalization outcome. This may have also reduced use of broader codes (i.e. those related to non-influenza-specific outcomes) in the hospital administrative database compared to before the COVID-19 era. These strengths increased the validity and reliability of our findings for the influenza hospitalizations outcomes, as laboratory-confirmed influenza hospitalization is generally the most specific vaccine effectiveness endpoint (26, 27).

Future research should focus on applying previously described techniques, such as PERR (Prior event rate ratio adjustment), to better adjust for confounding (28, 29). Additional research should also continue in subsequent years of HD-QIV use in France, though such research should proceed with caution as PCR testing practice in hospitals may regress post-COVID-19 pandemic, and broader hospitalization endpoints may continue be impacted by the observed “triple epidemic” of influenza, SARS-CoV-2 and RSV viruses co-circulating during winter; as observed during the 2022/23 season in France, causing a substantial burden on hospitals.

## Conclusion

HD-QIV was associated with a 23.3% (95%CI: 8.4–35.8) lower rate of influenza hospitalizations compared to SD-QIV. This is the first assessment of HD-QIV in France providing an overview of its performance in real-world setting. These findings are critical as they highlight the real-world reduction in severe clinical outcomes among older adults. The specificity of the influenza hospitalization endpoint is important in the context of this study (observational study, with observed residual confounding, and with high SARS-CoV-2 circulation). These findings, building on existing evidence across 10 influenza seasons in both randomized and observational studies, provide further evidence of the important clinical benefit of HD-QIV (6).

## Ethics approval and consent to participate

The data supporting the study findings are part of the National health data system (SNDS, Système national des Données de Santé) and are available from the HDH (Health Data Hub https://www.health-data-hub.fr/). Restrictions apply to the availability of these data containing potentially identifying and sensitive patient information. Special permission to access these data for this study was granted by the ethics and scientific committee for health research, studies, and evaluations (CESREES, Comité Ethique et Scientifique pour les Recherches, les Etudes et les Evaluations dans le domaine de la Santé) (former CEREES) and the French data protection authority (Comité National de l’Informatique et des Libertés, CNIL). The study protocol obtained two consecutive authorizations from the French data protection authority CNIL (initial authorization: Decision No. DR-2022-049; substantial modifications authorization: Decision DR-2023-013).

Informed consent was not required for the use of the anonymized secondary data, as mentioned in the Social Security Code, Article L161–28-1. All methods were performed in accordance with CNIL regulations and with REporting of studies Conducted using Observational Routinely-collected Data (RECORD) guidelines.

## Data Availability

The data that support the findings of this study are available from the French national health insurance information system, but restrictions apply to the availability of these data, which were used under license for the current study, and so are not publicly available.

## Acknowledgements

We would like to thank Marion Fournier and Marine Dufournet from Sanofi and Alexandre Vainchtock and Sacha Hiridjee from Heva for their strong contributions to this project, Rob Van Aalst and Matthew Loaicono for sharing methodological insights, and Fabrice Carrat who provided some comments to the protocol and statistical analysis plan. We thank the Direction de la Stratégie, des études et des statistiques (DSES), Département accès, traitements et analyse de la donnée (DATAD), and Cellule de la Cnam en Charge de l’accompagnement des Demandes D’extraction (DEMEX) teams at the Caisse Nationale de l’Assurance maladie (Cnam) for data extraction.

