## Supplementary for "The relative effectiveness of a high-dose quadrivalent influenza vaccine vs standard-dose quadrivalent influenza vaccines in older adults in France: a retrospective cohort study during the 2021-22 influenza season"

Bricout Hélène et al.

Table S1. Medication codes (UCD13 and CIP13) for SD-QIV and HD-QIV

| Vaccine | UCD13 code | CIP13 code |
| --- | --- | --- |
| Standard-dose QIV |  |  |
| - INFLUVAC TETRA SUSP INJ SER 0,5ML | 3400894338703 | 3400930117712 |
| - VAXIGRIPTETRA SUSP INJ SER 0,5ML | 3400894329657 | 3400928099877 |
|  | 3400894329657 | 3400930067727 |
| High-Dose QIV |  |  |
| - EFLUELDA SUSP INJ SER VACCIN | 3400890004602 | 3400930205372 |
|  |  | 3400930205389 |
|  |  | 3400930205426 |
|  |  | 3400930205402 |
|  |  | 3400930205396 |
|  |  | 3400930205419 |

Table S2. ICD-10 discharge codes for hospitalizations

| Definition | ICD-10 codes |
| --- | --- |
| Influenza hospitalisation | J09 to J11 |
| Pneumonia hospitalization | J12 to J18 |
| Respiratory hospitalization | J00 to J06, J09 to J18, J40-J41, J96 |
| Cardiovascular hospitalization | I16, I20 to I22, I24, I24, I26, I30, I40, I46 to I50, I63, I65-I66, G45-G46, J96 |
| Cardiorespiratory hospitalization | J00 to J06, J09 to J18, J40-J41, J96  I20 to I22, I24, I24, I26, I30, I40, I46 to I50, I63, I65-I66, G45-G46 |
| COVID-19 hospitalization | U071, U0710, U0711, U0714, U0715 |
| Urinary tract infection hospitalization | N410, N412-N413, N418, N419, N10, N110, N12, N136, N300, N309, T835 |
| Erysipelas hospitalization | A46 |
| Cataract hospitalization | BFGA427, BFGA004, BFPA002, BFGA368, BFGA008, BFGA002, BFGA003, BFGA010, BFGA006, GFGA009 |

Table S3 – ICD-10 codes or references to the codes used to identify comorbidities

| Comorbity | Codes or references |
| --- | --- |
| Diabetes | Rachas A, Gastaldi-Menager C, Denis P, Barthelemy P, Constantinou P, Drouin J, et al. The Economic Burden of Disease in France From the National Health Insurance Perspective: The Healthcare Expenditures and Conditions Mapping Used to Prepare the French Social Security Funding Act and the Public Health Act. Med Care. 2022;60(9):655-64. |
| Obesity and/or history of obesity surgery | HFCA001, HFCC003, HFFA001, HFFA011, HFFC004, HFFC018, HFGC900, HFKA001, HFKA002, HFKC001, HFLC900, HFLE002, HFMA009, HFMA010, HFMA011, HFMC006, HFMC007, HFMC008, HGCA009, HGCC027, E66 |
| Undernourishment/or history of undernourishment | E43, E44, E46 |
| COPD/Asthma | Rachas A, Gastaldi-Menager C, Denis P, Barthelemy P, Constantinou P, Drouin J, et al. The Economic Burden of Disease in France From the National Health Insurance Perspective: The Healthcare Expenditures and Conditions Mapping Used to Prepare the French Social Security Funding Act and the Public Health Act. Med Care. 2022;60(9):655-64. |
| Dementia | Rachas A, Gastaldi-Menager C, Denis P, Barthelemy P, Constantinou P, Drouin J, et al. The Economic Burden of Disease in France From the National Health Insurance Perspective: The Healthcare Expenditures and Conditions Mapping Used to Prepare the French Social Security Funding Act and the Public Health Act. Med Care. 2022;60(9):655-64. |
| Cardiovascular diseases | Rachas A, Gastaldi-Menager C, Denis P, Barthelemy P, Constantinou P, Drouin J, et al. The Economic Burden of Disease in France From the National Health Insurance Perspective: The Healthcare Expenditures and Conditions Mapping Used to Prepare the French Social Security Funding Act and the Public Health Act. Med Care. 2022;60(9):655-64. |
| Immunocompromised individuals | Rachas A, Gastaldi-Menager C, Denis P, Barthelemy P, Constantinou P, Drouin J, et al. The Economic Burden of Disease in France From the National Health Insurance Perspective: The Healthcare Expenditures and Conditions Mapping Used to Prepare the French Social Security Funding Act and the Public Health Act. Med Care. 2022;60(9):655-64.  And  Wyplosz B, Fernandes J, Goussiaume G, Moïsi J, Lortet-Tieulent J, Vainchtock A, et al. Adults at risk of pneumococcal disease in France. Infect Dis Now. 2021 Nov;51(8):661–6. |
| Chronic liver disease | Rachas A, Gastaldi-Menager C, Denis P, Barthelemy P, Constantinou P, Drouin J, et al. The Economic Burden of Disease in France From the National Health Insurance Perspective: The Healthcare Expenditures and Conditions Mapping Used to Prepare the French Social Security Funding Act and the Public Health Act. Med Care. 2022;60(9):655-64. |
| Terminal chronic kidney failure | Rachas A, Gastaldi-Menager C, Denis P, Barthelemy P, Constantinou P, Drouin J, et al. The Economic Burden of Disease in France From the National Health Insurance Perspective: The Healthcare Expenditures and Conditions Mapping Used to Prepare the French Social Security Funding Act and the Public Health Act. Med Care. 2022;60(9):655-64. |

Table S4 – Description of the characteristics of the unmatched HD-QIV community group

| **Characteristics** | **HD-QIV** |
| --- | --- |
| Number of individuals | 350 |
| Age, mean (± STD) | 85.1 (9.4) |
| - 65-74 years, n (%) | 66 (18.9) |
| - 75-85 years, n (%) | 93 (26.6) |
| - Over 85 years of age, n (%) | 191 (54.6) |
| Women, n (%) | 221 (63.1) |
| *Reasons for end of follow up, n (%)* |  |
| Admission into a medico-social housing (other than NH) | 0 (0.0) |
| Admission into NH | 1 (0.3) |
| Death | 20 (5.7) |
| End of follow-up | 329 (94.0) |
| Health care seeking behaviors proxy |  |
| All-cause hospitalization in the past 12 months, mean (STD) | 0.1 (0.3) |
| GP visits in the past 12 months, mean (STD) | 10.3 (18.0) |
| Influenza vaccination at pharmacy, n (%) | 167 (47.7) |
| Influenza vaccination during the previous season, n (%) | 290 (82.9) |
| COVID-19 vaccinated*, n (%) | 339 (96.9) |
| Pneumococcal vaccination in the previous 5 years, n (%) | 60 (17.1) |
| *Medical conditions during the 5 years prior index date, n (%)* |  |
| Diabetes | 72 (20.6) |
| Obesity and/or history of obesity surgery | 24 (6.9) |
| Undernourishment/or history of undernourishment | 81 (23.1) |
| COPD/Asthma | 41 (11.7) |
| Dementia | 109 (31.1) |
| Cardiovascular diseases | 127 (36.3) |
| Immunocompromised individuals | 67 (19.1) |
| Chronic liver disease | 11 (3.1) |
| Terminal chronic kidney failure | 2 (0.6) |
| *Number of comorbidities , n (%)* |  |
| None | 95 (27.1) |
| 1 | 120 (34.3) |
| 2 | 77 (22.0) |
| 3 | 33 (9.4) |
| 4 | 13 (3.7) |
| 5 | 7 (2.0) |
| 6 | 5 (1.4) |
| *Precariousness index (FDep99), n (%)* |  |
| Q1, Individuals living in a low-poverty municipality | 65 (18.6) |
| Q2 | 55 (15.7) |
| Q3 | 99 (28.3) |
| Q4 | 43 (12.3) |
| Q5, Individuals living in a highly disadvantaged municipality | 88 (25.1) |
| *COVID vaccinated is a variable identified as such within the database. It reflects the COVID-19 vaccination status of each patient at index date following current guidelines (it can refer to a single dose, two, or three, depending on the individual’s eligibility | |

Table S5: Baseline cohort characteristics (before matching procedure)

| **Characteristics** | **HD-QIV** | **SD-QIV** | **SMD** | **p-value** |
| --- | --- | --- | --- | --- |
| Number of individuals | 431,643 | 7,401,210 |  |  |
| Age, mean (± STD) | 77.4 (7.9) | 75.9 (7.7) | 0.1932 | <.0001 |
| - 65-74 years, n (%) | 182,449 (42.3) | 3,715,501 (50.2) | 0.0716 | <.0001 |
| - 75-85 years, n (%) | 170,954 (39.6) | 2,674,280 (36.1) | -0.1596 | <.0001 |
| - Over 85 years of age, n (%) | 78,240 (18.1) | 1,011,429 (13.7) | 0.1222 | <.0001 |
| Women, n (%) | 241,347 (55.9) | 4,028,165 (54.4) | 0.0299 | <.0001 |
| *Reasons for end of follow up, n (%)* | | |  |  |
| Admission into a medico-social housing (other than NH) | 35 (0.01) | 502 (0.01) | 0.0015 | <.0001 |
| Admission into NH | 1,454 (0.3) | 18,750 (0.3) | 0.0154 | <.0001 |
| Death | 8,526 (2.0) | 113,632 (1.5) | 0.0335 | <.0001 |
| End of follow-up | 421,628 (97.7) | 7,268,326 (98.2) | -0.037 | <.0001 |
| *Health care seeking behaviors proxy* | | |  |  |
| All-cause hospitalization in the past 12 months, mean (±STD) | 0.1 (±0.8) | 0.1 (±0.9) | -0.007 | <.0001 |
| GP visits in the past 12  months, mean (±STD) | 6.2 (±4.8) | 5.9 (±4.6) | 0.0462 | <.0001 |
| Influenza vaccination at pharmacy, n (%) | 217,949 (50.5) | 3,151,756 (42.6) | 0.1591 | <.0001 |
| Influenza vaccination during the previous season, n (%) | 394,207 (91.3) | 6,670,249 (90.1) | 0.0415 | <.0001 |
| COVID-19 vaccinated*, n (%) | 401,624 (93.1) | 6,925,111 (93.6) | -0.0209 | <.0001 |
| Pneumococcal vaccination in the previous 5 years, n (%) | 50,293 (11.7) | 840,538 (11.4) | 0.0092 | <.0001 |
| *Medical conditions during the 5 years prior index date, n (%)* | | |  |  |
| Diabetes | 85,249 (19.7) | 1,435,636 (19.4) | 0.0089 | <.0001 |
| Obesity and/or history of obesity surgery | 35,151 (8.1) | 583,339 (7.9) | 0.0096 | <.0001 |
| Undernourishment/or history of undernourishment | 28,524 (6.6) | 395,644 (5.4) | 0.0533 | <.0001 |
| COPD/Asthma | 50,644 (11.7) | 849,151 (11.5) | 0.0081 | <.0001 |
| Dementia | 15,519 (3.6) | 166,906 (2.3) | 0.0796 | <.0001 |
| Cardiovascular diseases | 119,671 (27.7) | 1,921,073 (26.0) | 0.0399 | <.0001 |
| Immunocompromised individuals | 79,214 (18.4) | 1,338,621 (18.1) | 0.0069 | <.0001 |
| Chronic liver disease | 6,767 (1.6) | 112,939 (1.5) | 0.0034 | 0.0296 |
| Terminal chronic kidney failure | 1,750 (0.4) | 30,833 (0.4) | -0.0017 | 0.2679 |
| *Number of chronic diseases, n (%)* | | |  | <.0001 |
| None | 195,208 (45.2) | 3,521,489 (47.6) | -0.0472 |  |
| 1 | 139,121 (32.2) | 2,320,849 (31.4) | 0.0187 |  |
| 2 | 62,279 (14.4) | 999,942 (13.5) | 0.0265 |  |
| 3 | 23,389 (5.4) | 377,056 (5.1) | 0.0145 |  |
| 4 | 8,185 (1.9) | 128,151 (1.7) | 0.0123 |  |
| 5 | 2,599 (0.6) | 39,723 (0.5) | 0.0087 |  |
| 6 | 862 (0.2) | 14,000 (0.2) | 0.0024 |  |
| *Precariousness index (FDep99), n (%)* |  |  |  | <.0001 |
| Q1, Individuals living in a low-poverty municipality | 78,066 (18.1) | 1,335,435 (18.0) | 0.0011 |  |
| Q2 | 79,508 (18.4) | 1,366,489 (18.5) | -0.0011 |  |
| Q3 | 85,262 (19.8) | 1,436,228 (19.4) | 0.0088 |  |
| Q4 | 79,365 (18.4) | 1,474,619 (19.9) | -0.0391 |  |
| Q5, Individuals living in a highly disadvantaged municipality | 83,567 (19.4) | 1,379,397 (18.6) | 0.0184 |  |
| Missing | 25,875 (6.0) | 409,042 (5.5) | 0.0201 |  |
| *COVID-19 vaccinated is a variable identified as such within the database. It reflects the COVID-19 vaccination status of each patient at index date following current guidelines (it can refer to a single dose, two, or three, depending on the individual’s eligibility and recommendation in place at index date)  SMD: Standardised Mean Differences | | | | |

Table S6 – Analysis with hospitalizations outcomes identified using ICD-10 primary or non-primary discharge codes, excluding COVID-19 codes

| **Hospitalization outcomes** | **HD-QIV event rate Per 100,000 person-years** | **SD-QIV event rate Per 100,000 person-years** | **IRR HD-QIV vs SD-QIV (95% CI)** |
| --- | --- | --- | --- |
| Influenza | 107.78 | 137.68 | 0.79 (0.68; 0.91) |
| Pneumonia | 1552.35 | 1480.66 | 1.05 (1.01; 1.10) |
| Pneumonia and influenza | 1644.98 | 1600.80 | 1.03 (0.99; 1.07) |
| Respiratory | 2871.45 | 2719.26 | 1.06 (1.03; 1.09) |
| Cardiovascular | 9687.57 | 9360.97 | 1.04 (1.02; 1.06) |
| Cardiorespiratory | 10377.2 | 10059.7 | 1.03 (1.01; 1.05) |

Table S7 – Analysis with hospitalizations outcomes identified using ICD-10 primary or non-primary discharge codes, including COVID-19 codes

| **Hospitalization outcomes** | **HD-QIV event rate Per 100,000 person-years** | **SD-QIV event rate Per 100,000 person-years** | **IRR HD-QIV vs SD-QIV (95% CI)** |
| --- | --- | --- | --- |
| Influenza | 112.42 | 144.19 | 0.78 (0.68; 0.90) |
| Pneumonia | 2,226.43 | 2,103.74 | 1.06 (1.03; 1.10) |
| Pneumonia and influenza | 2,338.84 | 2,247.93 | 1.05 (1.01; 1.08) |
| Respiratory | 3,654.57 | 3,452.71 | 1.06 (1.03; 1.10) |
| Cardiovascular | 10,412.6 | 10,057.8 | 1.04 (1.02; 1.06) |
| Cardiorespiratory | 14,067.2 | 13,510.5 | 1.05 (1.03; 1.07) |
